# COVID-19 vaccine response in pregnant and lactating women: a cohort study

**DOI:** 10.1101/2021.03.07.21253094

**Authors:** Kathryn J. Gray, Evan A. Bordt, Caroline Atyeo, Elizabeth Deriso, Babatunde Akinwunmi, Nicola Young, Aranxta Medina Baez, Lydia L. Shook, Dana Cvrk, Kaitlyn James, Rose M. De Guzman, Sara Brigida, Khady Diouf, Ilona Goldfarb, Lisa M. Bebell, Lael M. Yonker, Alessio Fasano, Sayed A. Rabi, Michal A. Elovitz, Galit Alter, Andrea G. Edlow

**Affiliations:** Department of Obstetrics and Gynecology, Brigham and Women’s Hospital, Harvard Medical School, Boston, MA, USA; Department of Pediatrics, Lurie Center for Autism, Massachusetts General Hospital, Harvard Medical School, Boston, MA, USA; Ragon Institute of MGH, MIT, and Harvard, Cambridge, MA, USA; PhD Program in Virology, Division of Medical Sciences, Harvard University, Boston, MA, USA; Department of Obstetrics and Gynecology, Massachusetts General Hospital, Harvard Medical School, Boston, MA, USA; Vincent Center for Reproductive Biology, Massachusetts General Hospital, Boston, MA, USA; Division of Infectious Diseases, Massachusetts General Hospital, MGH Center for Global Health, and Harvard Medical School, Boston, MA; Mucosal Immunology and Biology Research Center, Massachusetts General Hospital, Boston, MA; Department of Pediatrics, Massachusetts General Hospital, Boston, MA; Harvard Medical School, Boston, MA, USA; Department of Cardiothoracic Surgery, Massachusetts General Hospital, Harvard Medical School, Boston, MA, USA; Maternal and Child Health Research Center, Perelman School of Medicine, University of Pennsylvania, Philadelphia, PA, USA

**Keywords:** COVID-19 vaccine, mRNA, pregnancy, antibodies, breastmilk, breastfeeding, cord blood, maternal immunity, neonatal immunity

## Abstract

**Background:** Pregnant and lactating women were excluded from initial COVID-19 vaccine trials; thus, data to guide vaccine decision-making are lacking. We sought to evaluate the immunogenicity and reactogenicity of COVID-19 mRNA vaccination in pregnant and lactating women.

**Methods:** 131 reproductive-age vaccine recipients (84 pregnant, 31 lactating, and 16 non-pregnant) were enrolled in a prospective cohort study at two academic medical centers. Titers of SARS-CoV-2 Spike and RBD IgG, IgA and IgM were quantified in participant sera (N=131), umbilical cord sera (N=10), and breastmilk (N=31) at baseline, 2nd vaccine dose, 2-6 weeks post 2nd vaccine, and delivery by Luminex, and confirmed by ELISA. Titers were compared to pregnant women 4-12 weeks from native infection (N=37). Post-vaccination symptoms were assessed. Kruskal-Wallis tests and a mixed effects model, with correction for multiple comparisons, were used to assess differences between groups.

**Results:** Vaccine-induced immune responses were equivalent in pregnant and lactating vs non-pregnant women. All titers were higher than those induced by SARS-CoV-2 infection during pregnancy. Vaccine-generated antibodies were present in all umbilical cord blood and breastmilk samples. SARS-CoV-2 specific IgG, but not IgA, increased in maternal blood and breastmilk with vaccine boost. No differences were noted in reactogenicity across the groups.

**Conclusions:** COVID-19 mRNA vaccines generated robust humoral immunity in pregnant and lactating women, with immunogenicity and reactogenicity similar to that observed in non-pregnant women. Vaccine-induced immune responses were significantly greater than the response to natural infection. Immune transfer to neonates occurred via placental and breastmilk.

## INTRODUCTION

More than 71,000 infections and 79 maternal deaths have occurred in pregnant women in the United States alone as of February 2021^1^. SARS-CoV-2 infection is more severe in pregnant women compared to their non-pregnant counterparts, with an increased risk of hospital admission, ICU stay, and death^2^. Despite their higher risk, pregnant and lactating women were not included in any initial coronavirus disease 19 (COVID-19) vaccine trials, although the first vaccine trial began in pregnant women in February of 2021 (Pfizer/BioNTech, ClinicalTrials.gov Identifier: NCT04754594).

Pregnant women have long been left out of therapeutic and vaccine research reportedly due to heightened safety concerns in this population ^3–6^. Although the American College of Obstetricians and Gynecologists (ACOG) and the Society for Maternal-Fetal Medicine (SMFM) encouraged the Food and Drug Administration (FDA) to include pregnant women in the COVID-19 vaccine emergency use authorization (EUA) due to the risk of increased disease severity in this population, evidence about vaccine immunogenicity to guide patient decision-making and provider counseling is lacking^7–9^. Specifically, given the novelty of the first emergency approved COVID-19 vaccines, both of which utilize mRNA to deliver SARS-CoV-2 Spike to educate the immune system^10,11^, it remains unclear whether this novel vaccine approach will drive immunity in the context of pregnancy and location, and whether antibodies will be transferred efficiently to neonates via the cord and breastmilk. Additionally, characterizing vaccine side effects in pregnancy is important, particularly as fever is a common side effect reported and can pose potential risks in pregnancy. Here, vaccine-induced immunity was profiled in vaccinated pregnant, lactating and non –pregnant controls compared to women infected with SARS-CoV-2 during pregnancy. Robust immunity was observed across pregnant and lactating women, with detectable and potentially time-dependent, transfer of vaccine induced antibodies to neonates. Thus, mRNA vaccination results in robust vaccine-induced immunity during pregnancy that may provide critical protection to this vulnerable and unique population of mother:infant dyads.

## METHODS

### Study Design

Women at two tertiary care centers were approached for enrollment in an IRB-approved (protocol #2020P003538) COVID-19 pregnancy and lactation biorepository study between December 17, 2020 and February 23, 2021. Eligible women were: (1) pregnant; (2) lactating; or (3) non-pregnant and of reproductive age (18-45); ≥18 years old, able to provide informed consent, and receiving the COVID-19 vaccine.

### Participants and Procedures

Eligible study participants were identified by practitioners at the participating hospitals or were self-referred. A study questionnaire was administered to assess pregnancy and lactation status, history of prior SARS-CoV-2 infection, timing of COVID-19 vaccine doses, type of COVID-19 vaccine received (BNT162b2 Pfizer/BioNTech or mRNA-1273 Moderna/NIH), and side effects after each vaccine dose (injection site soreness, injection site skin reaction/rash, headache, myalgias, fatigue, fever/chills, allergic reaction, or other (reaction detailed). A cumulative symptom/reactogenicity score was generated by assigning one point to each side effect.

### Sample Collection and Processing

Blood and breastmilk from lactating women were collected at: V0 (at the time of first vaccine dose/baseline), V1 (at the time of second vaccine dose/”prime” profile), V2 (2-5.5 weeks following the 2nd vaccine dose/”boost” profile), and at delivery (for pregnant participants who delivered during the study timeframe). Umbilical cord blood was also collected at delivery for pregnant participants. The V2 timepoint reflects full antibody complement, achieved one week after Pfizer/BioNTech and two weeks after Moderna/NIH ^10,11^. Blood was collected by venipuncture (or from the umbilical vein following delivery for cord blood) into serum separator and EDTA tubes. Blood was centrifuged at 1000g for 10 min at room temperature. Sera and plasma were aliquoted into cryogenic vials and stored at-80°C.

### Antibody Quantification

Antibody quantification was performed as described previously ^12^. Briefly, a multiplexed Luminex assay was used to determine relative titer of antigen-specific isotypes and subclasses using the following antigens: SARS-CoV-2 Receptor Binding Domain (RBD), S1, and S2 (all Sino Biological), and SARS-CoV-2 Spike (LakePharma). Antigen-specific antibody titers were log10 transformed for time course analyses. PBS background intensity was reported for each antigen as a threshold for positivity. Titers resulting from natural infection and vaccination-induced antibodies against SARS-CoV-2 RBD and Spike were quantified from the same plate using ELISA as previously described^13^. Additional detail regarding antibody quantification may be found in Supplemental Methods.

### Statistical Analyses

Participant characteristics were summarized with frequency statistics. Continuous outcome measures were reported as either mean (standard deviation [SD]) or median (interquartile range [IQR]). Correlation analyses were performed using Spearman coefficients. Within and between group analyses of log10 transformed antibody levels in serum or breastmilk across multiple timepoints were evaluated by a repeated measures mixed effects (REML) model, followed by post-hoc Tukey’s multiple comparisons test. Differences between paired maternal and cord sera IgG were evaluated by Wilcoxon matched-pairs signed rank test. Statistical significance was defined as p<0.05. Statistical analyses were performed using GraphPad Prism 9 and Stata/IC version 16.1.

## RESULTS

From December 17, 2020 to March 2, 2021, samples were obtained from 131 enrolled participants: 84 pregnant, 31 lactating, and 16 non-pregnant reproductive-aged women. Of the pregnant vaccine recipients, 13 delivered during the study timeframe, and cord blood was collected at delivery from 10. Banked sera from 37 pregnant women infected with SARS-CoV-2 in pregnancy and enrolled between March 24, 2020 and December 11, 2020 were included as a second comparison group.

### Participant characteristics

Participant demographic and clinical characteristics, sampling timepoints, and side effect profiles are presented in Table 1. The study population consisted primarily of White, non-Hispanic women in their mid-30s, reflecting the healthcare worker population at the two hospitals. Five total participants reported prior SARS-CoV-2 infection: 2 pregnant, 2 lactating, 1 non-pregnant. Characteristics of the comparison group with natural SARS-CoV-2 infection in pregnancy are detailed in Table S1.

**Table 1.**
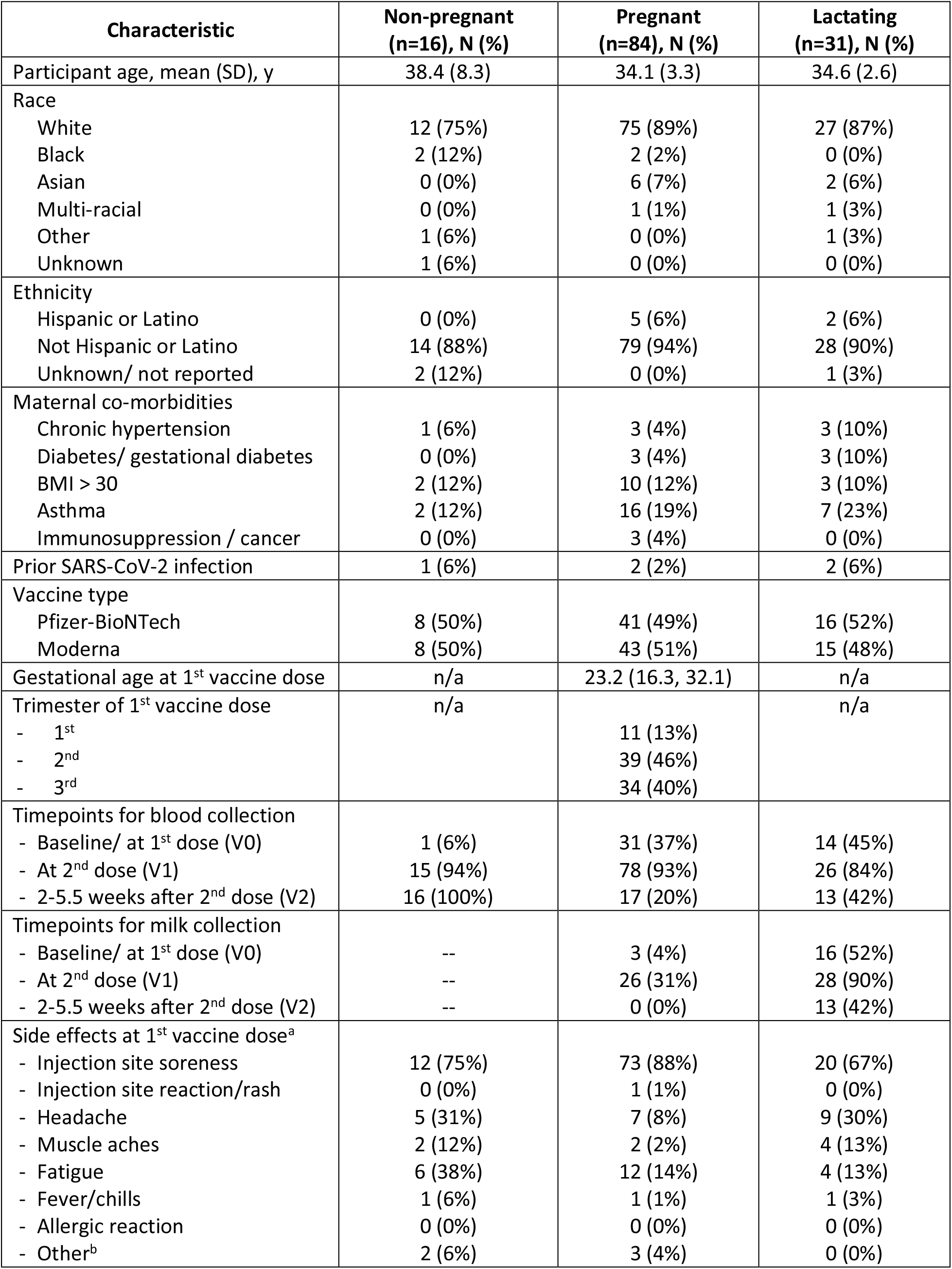
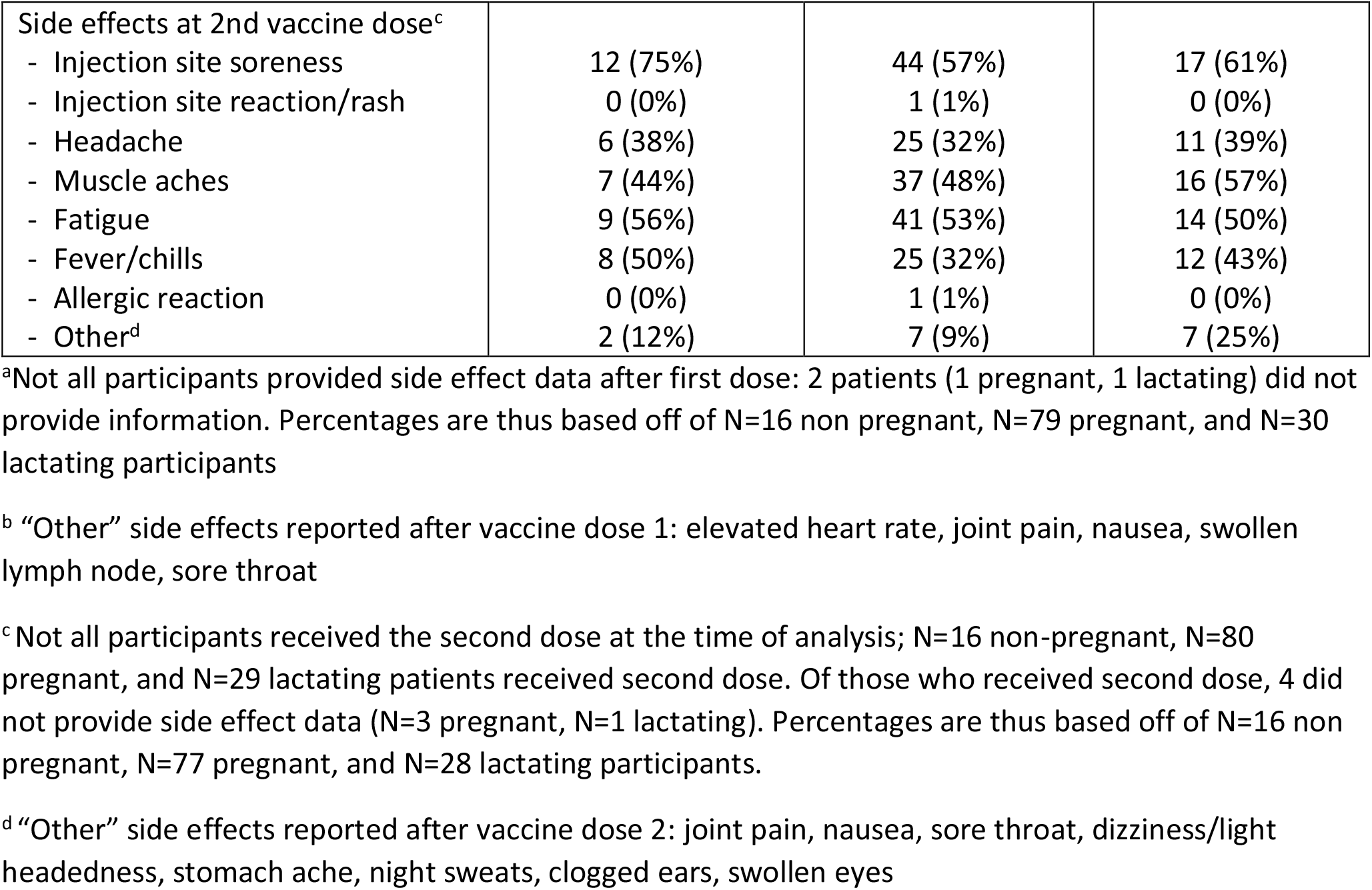
Cohort Demographic Characteristics.

| **Characteristic** | **Non-pregnant**<br>**(n=16), N (%)** | **Pregnant**<br>**(n=84), N (%)** | **Lactating**<br>**(n=31), N (%)** |
| --- | --- | --- | --- |
| Participant age, mean (SD), y | 38.4 (8.3) | 34.1 (3.3) | 34.6 (2.6) |
| Race |  |  |  |
| White | 12 (75%) | 75 (89%) | 27 (87%) |
| Black | 2 (12%) | 2 (2%) | 0 (0%) |
| Asian | 0 (0%) | 6 (7%) | 2 (6%) |
| Multi-racial | 0 (0%) | 1 (1%) | 1 (3%) |
| Other | 1 (6%) | 0 (0%) | 1 (3%) |
| Unknown | 1 (6%) | 0 (0%) | 0 (0%) |
| Ethnicity |  |  |  |
| Hispanic or Latino | 0 (0%) | 5 (6%) | 2 (6%) |
| Not Hispanic or Latino | 14 (88%) | 79 (94%) | 28 (90%) |
| Unknown/ not reported | 2 (12%) | 0 (0%) | 1 (3%) |
| Maternal co-morbidities |  |  |  |
| Chronic hypertension | 1 (6%) | 3 (4%) | 3 (10%) |
| Diabetes/ gestational diabetes | 0 (0%) | 3 (4%) | 3 (10%) |
| BMI > 30 | 2 (12%) | 10 (12%) | 3 (10%) |
| Asthma | 2 (12%) | 16 (19%) | 7 (23%) |
| Immunosuppression / cancer | 0 (0%) | 3 (4%) | 0 (0%) |
| Prior SARS-CoV-2 infection | 1 (6%) | 2 (2%) | 2 (6%) |
| Vaccine type |  |  |  |
| Pfizer-BioNTech | 8 (50%) | 41 (49%) | 16 (52%) |
| Moderna | 8 (50%) | 43 (51%) | 15 (48%) |
| Gestational age at 1<sup>st</sup> vaccine dose | n/a | 23.2 (16.3, 32.1) | n/a |
| Trimester of 1<sup>st</sup> vaccine dose | n/a |  | n/a |
| - 1<sup>st</sup> |  | 11 (13%) |  |
| - 2<sup>nd</sup> |  | 39 (46%) |  |
| - 3<sup>rd</sup> |  | 34 (40%) |  |
| Timepoints for blood collection |  |  |  |
| - Baseline/ at 1<sup>st</sup> dose (V0) | 1 (6%) | 31 (37%) | 14 (45%) |
| - At 2<sup>nd</sup> dose (V1) | 15 (94%) | 78 (93%) | 26 (84%) |
| - 2-5.5 weeks after 2<sup>nd</sup> dose (V2) | 16 (100%) | 17 (20%) | 13 (42%) |
| Timepoints for milk collection |  |  |  |
| - Baseline/ at 1<sup>st</sup> dose (V0) | -- | 3 (4%) | 16 (52%) |
| - At 2<sup>nd</sup> dose (V1) | -- | 26 (31%) | 28 (90%) |
| - 2-5.5 weeks after 2<sup>nd</sup> dose (V2) | -- | 0 (0%) | 13 (42%) |
| Side effects at 1<sup>st</sup> vaccine dose<sup>a</sup> |  |  |  |
| - Injection site soreness | 12 (75%) | 73 (88%) | 20 (67%) |
| - Injection site reaction/rash | 0 (0%) | 1 (1%) | 0 (0%) |
| - Headache | 5 (31%) | 7 (8%) | 9 (30%) |
| - Muscle aches | 2 (12%) | 2 (2%) | 4 (13%) |
| - Fatigue | 6 (38%) | 12 (14%) | 4 (13%) |
| - Fever/chills | 1 (6%) | 1 (1%) | 1 (3%) |
| - Allergic reaction | 0 (0%) | 0 (0%) | 0 (0%) |
| - Other<sup>b</sup> | 2 (6%) | 3 (4%) | 0 (0%) |
| Side effects at 2nd vaccine dose<sup>c</sup> |  |  |  |
| - Injection site soreness | 12 (75%) | 44 (57%) | 17 (61%) |
| - Injection site reaction/rash | 0 (0%) | 1 (1%) | 0 (0%) |
| - Headache | 6 (38%) | 25 (32%) | 11 (39%) |
| - Muscle aches | 7 (44%) | 37 (48%) | 16 (57%) |
| - Fatigue | 9 (56%) | 41 (53%) | 14 (50%) |
| - Fever/chills | 8 (50%) | 25 (32%) | 12 (43%) |
| - Allergic reaction | 0 (0%) | 1 (1%) | 0 (0%) |
| - Other<sup>d</sup> | 2 (12%) | 7 (9%) | 7 (25%) |

<sup>a</sup>Not all participants provided side effect data after first dose: 2 patients (1 pregnant, 1 lactating) did not provide information. Percentages are thus based off of N=16 non pregnant, N=79 pregnant, and N=30 lactating participants
<sup>b</sup> “Other” side effects reported after vaccine dose 1: elevated heart rate, joint pain, nausea, swollen lymph node, sore throat
<sup>c</sup> Not all participants received the second dose at the time of analysis; N=16 non-pregnant, N=80 pregnant, and N=29 lactating patients received second dose. Of those who received second dose, 4 did not provide side effect data (N=3 pregnant, N=1 lactating). Percentages are thus based off of N=16 non pregnant, N=77 pregnant, and N=28 lactating participants.
<sup>d</sup> “Other” side effects reported after vaccine dose 2: joint pain, nausea, sore throat, dizziness/light headedness, stomach ache, night sweats, clogged ears, swollen eyes

### Vaccination characteristics

At the time of the study, two COVID-19 vaccines had received EUA: Pfizer/BioNTech and Moderna. Both vaccines use mRNA to deliver the SARS-CoV-2 Spike antigen to the immune system^10,11^, representing a novel vaccine platform never before tested in pregnancy. While mRNA vaccines have shown highly effective immune induction in non-pregnant adults, the immunogenicity and reactogenicity of this platform in pregnancy remains unclear. Equivalent numbers of pregnant women receiving the Pfizer/BioNTech and Moderna vaccines were included in our study. Of pregnant participants, the mean gestational age at first vaccine dose was 23.2 weeks, with 11 women (13%) receiving their first vaccine dose in the first trimester, 39 (46%) in second trimester, and 34 (40%) in the third trimester. Side effect profiles between participant groups following vaccination were similar. Injection site soreness, fatigue, and headache were the most common side effects after each vaccine dose, while fever/chills were also common after the 2^nd^ dose (Table 1). The cumulative symptom score after the 1 ^st^ dose in all three groups was low. After the 2^nd^ dose, there was no significant difference between groups with respect to cumulative symptom score (median (IQR) 2 (1-3), 3 (2-4), and 2.5 (1-4.5) in pregnant, lactating, and non-pregnant groups respectively, p = 0.40). Vaccine-related fevers/chills were reported by 32% (25/77) of pregnant women after the boost dose and 50% (8/16) of non-pregnant (p=0.25).

### Delivery outcomes and characteristics of lactating women

Delivery information for the 13 pregnant participants who delivered during the study period is detailed in Table 2. All 13 were vaccinated in the third trimester. Three women delivered at hospitals other than the study sites and cord blood samples were not available. Of the ten umbilical cord blood samples available for analysis, 9/10 mothers had received both vaccine doses (median (IQR) 36.5 days (30-42) from vaccine 1 and 14 days (11-16) from vaccine 2). One participant delivered 17 days after vaccine 1, with spontaneous preterm labor at 35 weeks’ gestation. Lactating participant characteristics are detailed in Table 2.

**Table 2.**
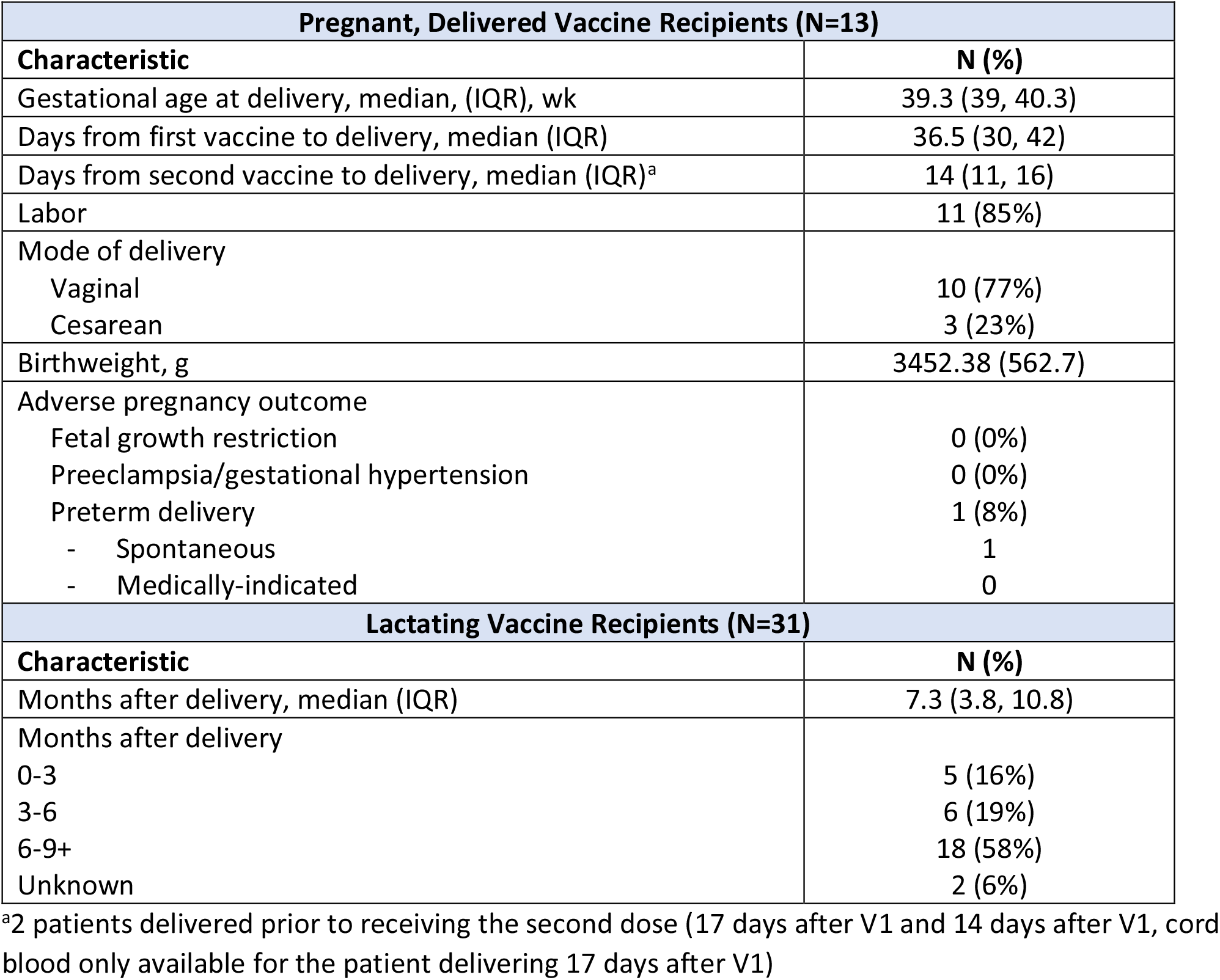
Characteristics of Pregnant, Delivered Vaccine Recipients and Lactating Vaccine Recipients.

| **Pregnant, Delivered Vaccine Recipients (N=13)** |  |
| --- | --- |
| **Characteristic** | **N (%)** |
| Gestational age at delivery, median, (IQR), wk | 39.3 (39, 40.3) |
| Days from first vaccine to delivery, median (IQR) | 36.5 (30, 42) |
| Days from second vaccine to delivery, median (IQR)<sup>a</sup> | 14 (11, 16) |
| Labor | 11 (85%) |
| Mode of delivery |  |
| Vaginal | 10 (77%) |
| Cesarean | 3 (23%) |
| Birthweight, g | 3452.38 (562.7) |
| Adverse pregnancy outcome |  |
| Fetal growth restriction | 0 (0%) |
| Preeclampsia/gestational hypertension | 0 (0%) |
| Preterm delivery | 1 (8%) |
| - Spontaneous | 1 |
| - Medically-indicated | 0 |
| **Lactating Vaccine Recipients (N=31)** |  |
| **Characteristic** | **N (%)** |
| Months after delivery, median (IQR) | 7.3 (3.8, 10.8) |
| Months after delivery |  |
| 0-3 | 5 (16%) |
| 3-6 | 6 (19%) |
| 6-9+ | 18 (58%) |
| Unknown | 2 (6%) |

<sup>a</sup>2 patients delivered prior to receiving the second dose (17 days after V1 and 14 days after V1, cord blood only available for the patient delivering 17 days after V1)

### The maternal vaccine response

IgM, IgG, and IgA responses to the Spike (S), receptor binding domain (RBD), S1-segment of S, and S2-segment of S were measured. A significant rise in all isotypes across all antigens was observed from V0 to V1, with a further rise in IgG levels from V1 to V2 in both the pregnant and lactating groups (**Fig 1A-D and Supplemental Fig 1**). Spike titers rose more rapidly than RBD-titers after the first (V1/prime timepoint) and second (V2/boost timepoint) vaccine dose, but the magnitude of the response did not differ across pregnant or lactating women. In contrast to IgG responses, IgM and IgA responses were induced robustly after the prime, and were poorly induced after boosting, across all groups (**Fig 1C and D**). Higher S-and RBD-specific IgA responses were noted in Moderna vaccinees compared to Pfizer/BioNTech vaccinees (**Supplemental Fig 2A-C**), potentially related to the extended boosting window used for the Moderna vaccine. By 2 weeks post-second vaccine, the dominant serum antibody response was IgG for pregnant, lactating, and non-pregnant women (**Fig 1E and Supplemental Fig 1C**). Strikingly higher levels of SARS-CoV-2 antibodies were observed in all vaccinated women compared to pregnant women with natural infection 4-12 weeks prior (**Fig 1F**, Kruskal Wallis p<0.001), highlighting the robust humoral immune responses induced by mRNA vaccination.

**Figure 1.**
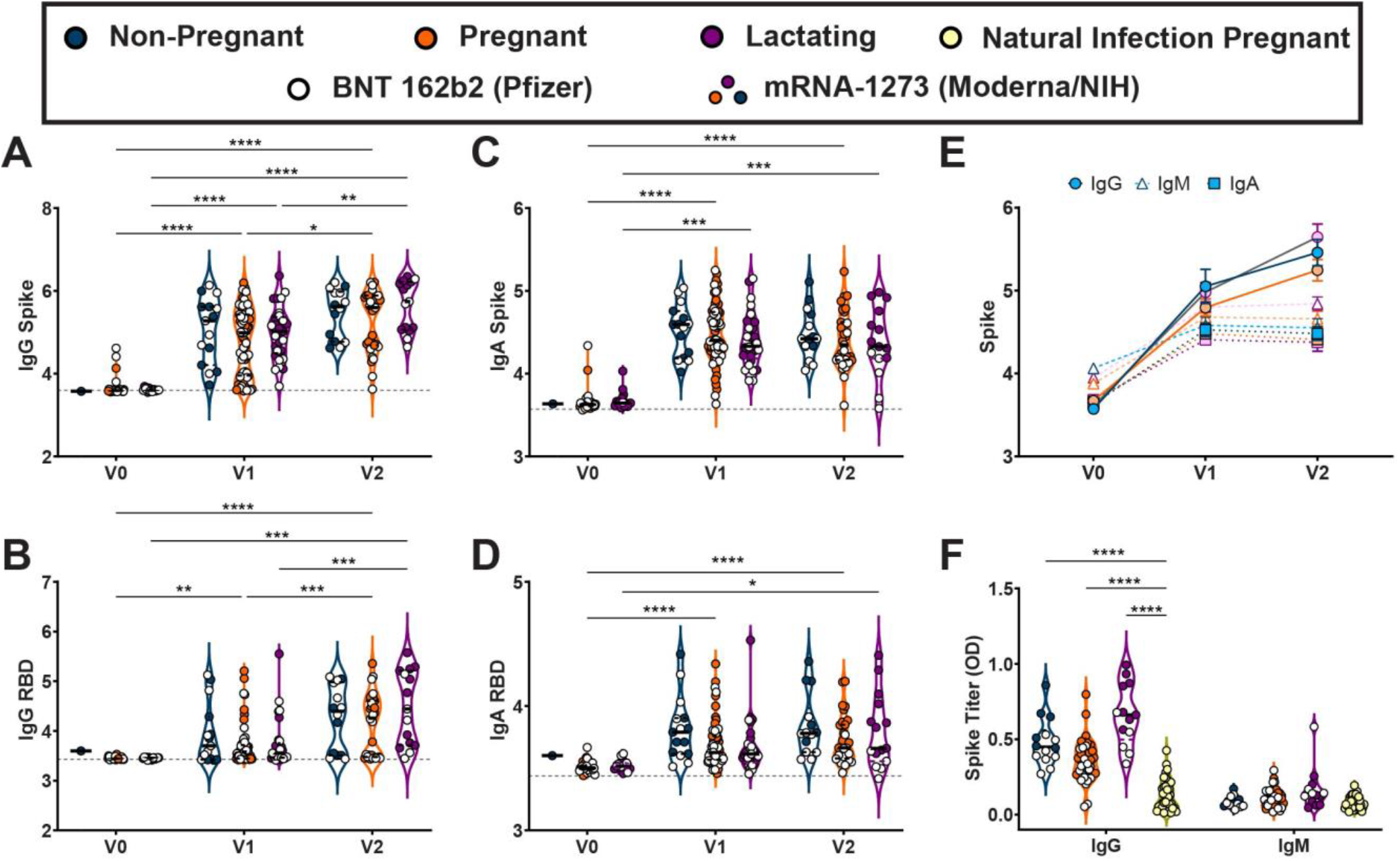
Maternal vaccination induces a robust SARS-CoV-2-specific antibody response. **A-D**. Violin plots show the Iog10 transformed **(A)** IgG Spike-, **(B)** IgG RBD-, **(C)** IgA Spike-, and **(D)** IgA RBD-specific titers across VO, V1, and V2 time points collected from non-pregnant reproductive-age (blue), pregnant (orange), or lactating (purple) participants. Participants who received BNT 162b2 from Pfizer/BioNTech are depicted as open circles, and participants who received mRNA-1273 from Moderna-/NIH are depicted as closed circles. Differences across timepoints and groups were assessed by repeated measures mixed-effects model followed by posthoc Tukey’s multiple comparisons test. * p <0.05, ** p <0.01, *** p <0.001, **** p <0.0001. **E**. Line graph showing the log 10 transformed relative Spike-specific titers across V0, V1, and V2 time points collected from non-pregnant (blue), pregnant (orange), or lactating (purple) participants for IgG (circles:solid lines), IgM (open triangles:dashed lines), and IgA (squares:dotted lines). **F**. Violin plots show the IgG and IgM Spike-specific titer in non-pregnant (blue), pregnant (orange), lactating (purple), and naturally-infected pregnant (yellow) participants. Participants who received BNT 162b2 from Pfizer/BioNTech are depicted as open circles, and participants who received mRNA-1273 from Moderna/NIH are depicted as closed circles. Differences across groups were assessed by Kruskal-Wallis test followed by posthoc Dunn’s multiple comparisons test. **** p <0.0001 compared to natural infection in pregnant women.

### Impact of maternal vaccination on breastmilk antibody transfer

mRNA vaccination resulted in the induction of antibodies in the circulation of vaccinated women (**Fig 1**). However, whether these antibodies were transferred efficiently to infants remained unclear. Thus, we next examined the levels of antibodies in breastmilk of lactating mothers (**Fig 2 A-C**). Robust induction of IgG, IgA, and IgM were observed following the prime and boost. Interestingly, IgA and IgM levels did not increase with boosting, in synchrony with a minimal boost in these isotypes in serum (**Fig 1C/D and Supplemental Fig 1A-E**). However, a boost in breastmilk IgG levels was observed (**Fig 2A)**, concomitant with the boost observed systemically/in maternal serum (**Fig 1A**). IgG1 RBD rose significantly from V0 to V2 (3.44 to 3.50, p = 0.002) but not V0 to V1 (3.44 to 3.45, p=0.7) in breastmilk, and there was no significant rise in anti-RBD IgA or IgM in breastmilk after either dose (**Supplemental Fig 3**). Overall these data suggest that the boost may drive enhanced breastmilk-transfer of IgG, in the setting of consistent unboosted IgA transfer.

**Figure 2.**
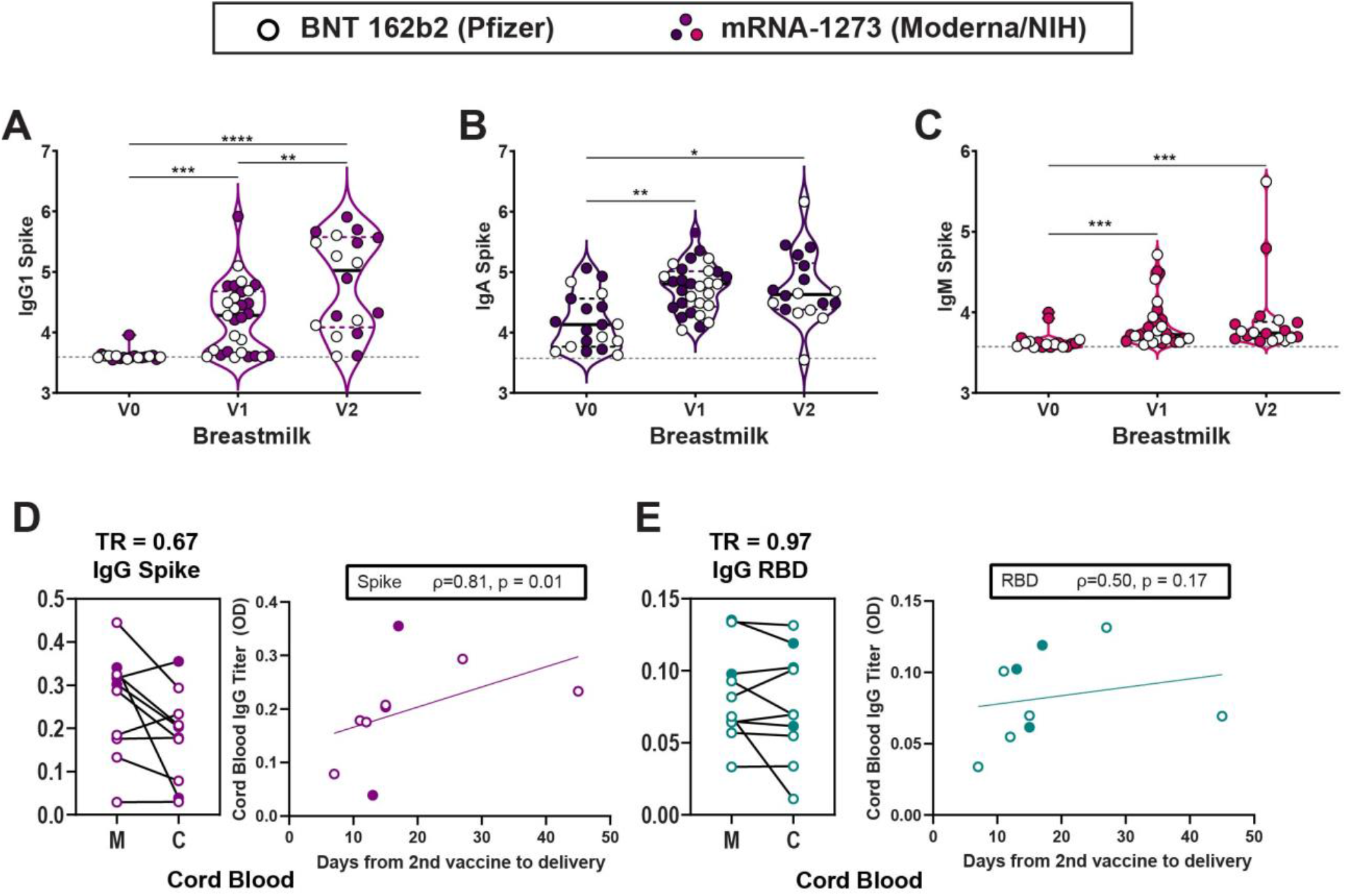
Maternal vaccination induces SARS-CoV-2-specific antibodies that transfer to breastmilk and umbilical cord blood. **A-C**. Violin plots show the Iog10 transformed **(A)** lgG1, **(B)** IgA, and **(C)** IgM Spike-specific breastmilk titers across VO, V1, and V2 time points. Differences across timepoints were assessed with repeated measures mixed effects model followed by posthoc Tukey’s multiple comparisons test. Participants who received BNT 162b2 from Pfizer/BioNTech are depicted as open circles, and participants who received mRNA-1273 from Moderna/NIH are depicted as closed circles. * p <0.05, ** p <0.01, *** p <0.001, **** p <0.0001. **D-E**. Dot plots showing relative (**D**) Spike-and (**E**) RBD-specific maternal blood (M) and cord blood (C) titers of lgG1. Wilcoxon matched-pairs signed rank test was performed to determine significance. TR = transfer ratio (median). On the right of each panel, the x-axis shows the time from 2nd vaccine until delivery and the y axis shows cord blood log 10 transformed titer for (**D**) IgG Spike (purple:solid line) and (**E**) IgG RBD (turquoise:dashed line). Correlation was determined by Spearman correlation test. PBS background subtraction was used to determine corrected optical density of 0.0.

### Impact of maternal vaccination on placental antibody transfer

Maternal IgG is also capable of crossing the placenta to confer immunity to the neonate. Spike-and RBD-specific IgG were detectable in 10/10 umbilical cords after maternal vaccination (**Fig 2D/E**). The cord with the lowest Spike- and RBD-specific IgG belonged to a mother who delivered between the first and second vaccine doses and had received her first vaccine dose 17 days prior to delivery, suggesting that 2 doses may be essential to optimize humoral immune transfer to the neonate. Interestingly, there was a significant improvement of transfer of S-, but not RBD-, specific IgG1 into the cord with time from boost (**Fig 2D/E**), suggesting that time from vaccination may be an important determinant of transfer rates of specific IgG subpopulations following immunization in pregnancy (**Supplemental Fig 4A/B)**.

### Vaccine reactogenicity in pregnancy and lactation

Composite reactogenicity score after boost dose of vaccine was significantly positively correlated with both maternal serum and breastmilk antibody titers. Composite symptom score after vaccination was significantly positively correlated with maternal serum Spike- and RBD-specific IgG1 and IgG3, breastmilk anti-Spike IgG1, IgG3 and IgA, and breastmilk anti-RBD IgG1 (Table S2). Within the pregnant women, medical comorbidities were not significantly associated with maternal serum antibody titers, although there were relatively few medical comorbidities in this group.

## DISCUSSION

The COVID-19 pandemic has given rise to an explosion of hundreds of vaccine platforms that are in development to fight SARS-CoV-2^14,15^. However, few of these platforms have been tested or are specifically designed to elicit immunity in vulnerable populations, including pregnant women. Due to the heightened susceptibility of pregnant women to severe COVID-19 disease, obstetrician/gynecologists have advocated for the importance of providing pregnant women and their infants with immunity ^7–9^. Critically, pregnant women were not included in COVID-19 vaccine clinical trials, and in fact, few vaccine platforms have been profiled in pregnant women, due to enhanced safety concerns in this population related to potential maternal, fetal, or obstetric morbidity^3–6^. Whether the first two EUA-approved vaccines, both of which exploit a novel mRNA-based vaccine strategy, could elicit humoral immunity in pregnant and lactating women was unclear. Here, robust and comparable IgG titers were observed across pregnant, lactating, and non-pregnant controls, all of which were significantly higher than those observed in pregnant women with prior SARS-CoV-2-infection. Boosting resulted in augmented IgG levels in the blood, translating to transfer of IgG to the neonate through the placenta and breastmilk.

The lack of boosting of IgM was likely related to an expected class switching to IgG, observed with increasing IgG titers observed following the boost. Conversely, the lack of boosting of IgA observed across all women in this study was unexpected. This lack of IgA augmentation may be related to the intramuscular administration of the vaccine, which triggers a robust induction of systemic, but not mucosal, antibodies. However, higher levels of IgA were noted after the boost in pregnant Moderna recipients, potentially attributable to enhanced class switching following a longer boosting interval. Robust IgG levels were noted in all vaccinees, and vaccine-induced IgG was transferred across the placenta to the fetus, as has been noted in the setting of influenza, pertussis, and other vaccination in pregnancy ^16–18^. Additionally, while the transferred levels of IgA through breastmilk did not increase with boosting, IgG transfer increased significantly with boost, resulting in the delivery of high levels of IgG to the neonate through breastmilk. Importantly, emerging data point to a critical role for breastmilk IgG in neonatal immunity against several other vaccinatable viral pathogens including HIV, RSV, and influenza ^19–21^. In contrast, IgA dominates breastmilk profiles in natural SARS-CoV-2 infection^22^. The different isotype transfer profile for breastmilk (IgG in vaccine, IgA in natural infection) likely reflects differences in antibody profile programming across mucosally-acquired natural SARS-CoV-2 infection versus intra-muscular vaccination. Whether breastmilk IgG or IgA will be more critical for neonatal protection remains unclear.

Based on what is known about other vaccines, the amount of maternal IgG transferred across the placenta to the cord is likely to differ by trimester of vaccination ^16,17^. Understanding transfer kinetics across all pregnancy trimesters will be an important direction for future research. While timing maternal COVID-19 vaccination may not be possible during this phase of the pandemic, understanding optimal timing of vaccination to augment neonatal humoral immunity remains important. Unlike vaccines that aim to boost pre-exiting antibodies, including the influenza and pertussis vaccines, optimal timing for de novo vaccine administration remains unclear. Thus, as the prevalence of SARS-CoV-2 community spread decreases, different factors such as optimizing neonatal immunity via placental or breastmilk transfer may be weighted more heavily to inform future vaccine deployment.

Following EUA for the COVID-19 mRNA vaccines, safety information has been tracked by the CDC using the V-safe smartphone application. Consistent with our observations, the V-safe data indicate no significant differences in post-vaccination reactions in pregnant vs. non-pregnant women aged 16-54 years^23^. While the side effect profile of pregnant women receiving the COVID-19 vaccines was not significantly different from non-pregnant women, the relatively high incidence of fever (up to 32% following the second dose), raises a theoretical concern for pregnant recipients. Febrile morbidity in pregnancy has been associated with congenital malformations ^24^ when it occurs in the first trimester, and adverse neurodevelopmental outcomes in offspring ^25^, although the level of risk remains controversial ^26^.

When considering vaccination in pregnancy, evidence regarding maternal and fetal benefit, as well as potential maternal and fetal harm and effects on pregnancy outcomes should be weighed carefully. While the absolute risk of severe COVID-19 is low in pregnant women, pregnancy is a risk factor for severe disease^27,28.^There are well-documented maternal, neonatal, and obstetric risks of SARS-CoV-2 infection during pregnancy ^29–33^. These data provide a compelling argument that COVID-19 mRNA vaccines induce similar humoral immunity in pregnant and lactating women as in the non-pregnant population. These data do not elucidate potential risks to the fetus. Thus, future studies, in larger populations, across gestational ages may enhance our ability to develop evidence-based recommendations for the administration of vaccines, and particularly different platforms, during pregnancy.

COVID-19 vaccination in pregnancy and lactation generated robust humoral immunity similar to that observed in non-pregnant women with similar side effect profiles. While humoral immune response and side effects are only two of many considerations for pregnant women and their care providers in weighing whether or not to be vaccinated against COVID-19 in pregnancy, these data confirm that the COVID19 mRNA vaccines result in comparable humoral immune responses in pregnant and lactating women to those observed in non-pregnant populations.

## Data Availability

All data referred to in the manuscript are available upon request.

