## Supplementary Materials for "COVID-19 vaccine response in pregnant and lactating women: a cohort study"

**SUPPLEMENTARY APPENDIX**

**Table of Contents**


| | |
|---|---|
| Supplemental Methods | page 2 |
| Supplemental Figures | pages 3-6 |
| Supplemental Figure 1. Maternal vaccination efficiently induces SARS-CoV-2-specific antibodies in maternal serum. | page 3 |
| Supplemental Figure 2. mRNA-1273 induces a greater IgA response than does BNT 162b2. | page 4 |
| Supplemental Figure 3. Maternal vaccination induces SARS-CoV-2-specific antibodies in breastmilk. | page 5 |
| Supplemental Figure 4. Transfer of SARS-CoV-2-specific antibodies from maternal to umbilical cord blood following maternal vaccination. | page 6 |
| Supplemental Tables | pages 7-8 |
| Table S1. Characteristics of pregnant women with natural COVID-19 infection | page 7 |
| Table S2. SARS-CoV-2-specific antibody titer correlation with composite participant symptom score after vaccine dose 2. | page 8 |
| Supplemental References | page 9 |


---

**TITLE:** COVID-19 vaccine response in pregnant and lactating women: a cohort study

**AUTHORS**: Kathryn J. Gray, MD PhD, Evan A. Bordt, PhD, Caroline Atyeo, BS, Elizabeth Deriso, PhD, Babatunde Akinwunmi, MD MPH MMSc, Nicola Young, BA, Aranxta Medina Baez, BS, Lydia L. Shook, MD, Dana Cvrk, CNM, Kaitlyn James, PhD, MPH, Rose M. De Guzman, PhD, Sara Brigida, BA, Khady Diouf, MD, Ilona Goldfarb, MD MPH, Lisa M. Bebell, MD, Lael M.Yonker, MD, Alessio Fasano, MD, Sayed A. Rabi, MD, Michal A. Elovitz, MD, Galit Alter, PhD, Andrea G. Edlow, MD, MSc

### Supplemental Methods

#### *Luminex-based antibody quantification*

Antibody quantification was performed as described previously[1]**.** Briefly, a multiplexed Luminex assay was used to determine relative titer of antigen-specific isotypes and subclasses using the following antigens: SARS-CoV-2 Receptor Binding Domain (RBD), S1, S2 (all Sino Biological) and SARS-CoV-2 Spike (LakePharma). Antigens were covalently linked to carboxyl-modified Magplex© Luminex beads using Sulfo-NHS (N-hydroxysulfosuccinimide, Pierce) and ethyl dimethylaminopropyl carbodiimide hydrochloride (EDC). Antigen-coupled microspheres were blocked, washed, resuspended in PBS, and stored at 4°C.

To form immune complexes, appropriately diluted plasma (1:100 for IgG2/3, IgA1, IgM; 1:500 for IgG1) or breastmilk (1:5 for IgG1, IgA1, and IgM), was added to the antigen-coupled microspheres, and plates were incubated overnight at 4˚C, shaking at 700 rpm. The following day, plates were washed with 0.1% BSA 0.02% Tween-20. PE-coupled mouse anti-human detection antibodies (Southern Biotech) were used to detect antigen-specific antibody binding. Fluorescence was acquired using an Intellicyt iQue, and relative antigen-specific antibody titer was log10 transformed for time course blood and breastmilk analyses. PBS background intensity was reported for each antigen as a threshold for positivity.

#### *Antibody Quantification Using Enzyme-linked Immunosorbent Assay (ELISA)*

Antibodies against SARS-CoV-2 RBD and Spike, were quantified using ELISA as previously described[2]. Briefly, plates were coated with 500 ng/mL per well of SARS-CoV-2 RBD, SARS-CoV-2 Spike, or SARS-CoV-2 N. Plates were incubated for 30 minutes at room temperature and washed in wash buffer (0.05% Tween-20, 400 mM NaCl, 50 mM Tris, pH 8.0). Plates were blocked with a 1% BSA solution then washed again with wash buffer. Serum samples were diluted 1:100 and added to the plates. Plates were incubated at 37°C for 30 minutes. After incubation, plates were washed, and anti-human IgG or anti-human IgM coupled to horseradish peroxidase (HRP) (Bethyl Laboratories) was added for detection. Plates were incubated for 30 minutes at room temperature and washed. The ELISA was developed with TMB and stopped with sulfuric acid. The signal was read at 450 nm and background corrected from a reference wavelength of 570 nm.

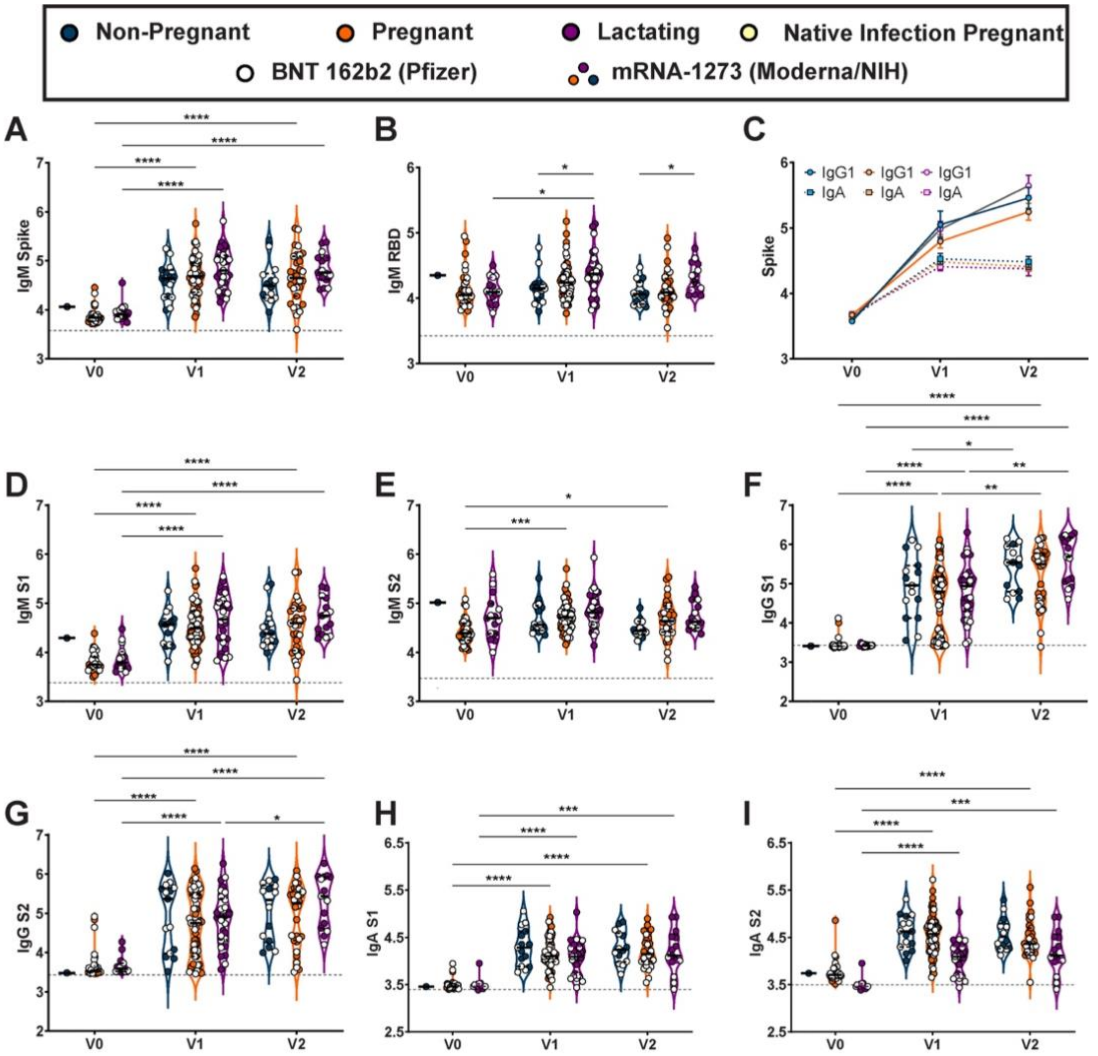


**Supplemental Figure 1. Maternal vaccination induces robust SARS-CoV-2-specific antibodies in maternal serum.**

**A-B.** Violin plots show the log10 transformed **(A)** IgM Spike- and **(B)** IgM RBD-specific titers across V0, V1, and V2 time points collected from non-pregnant controls (blue), pregnant (orange), or lactating (purple) patients. Participants injected with BNT 162b2 from Pfizer/BioNTech are depicted as open circles, and participants injected with mRNA-1273 from Moderna/NIH are depicted as closed circles. Differences across timepoints and groups were assessed by repeated measures mixed-effects model followed by posthoc Tukey's multiple comparisons test. * $p < 0.05$, ** $p < 0.01$, *** $p < 0.001$, **** $p < 0.0001$.

**C.** Line graph showing the log10 transformed relative Spike-specific titers across V0, V1, and V2 time points collected from non-pregnant controls (blue), pregnant (orange), or lactating (purple) patients for IgG (circles:solid lines) and IgA (squares:dotted lines).

**D-I.** Violin plots show the log10 transformed **(D)** IgM S1-, **(E)** IgM S2-, **(F)** IgG S1-, **(G)** IgG S2-, **(H)** IgA S1-, and **(I)** IgA S2-specific titers across V0, V1, and V2 time points collected from non-pregnant controls (blue), pregnant (orange), or lactating (purple) patients. Participants injected with BNT 162b2 from Pfizer/BioNTech are depicted as open circles, and participants injected with mRNA-1273 from Moderna/NIH are depicted as closed circles. Differences across timepoints and groups were assessed by repeated measures mixed-effects model followed by posthoc Tukey's multiple comparisons test. * $p < 0.05$, ** $p < 0.01$, *** $p < 0.001$, **** $p < 0.0001$.

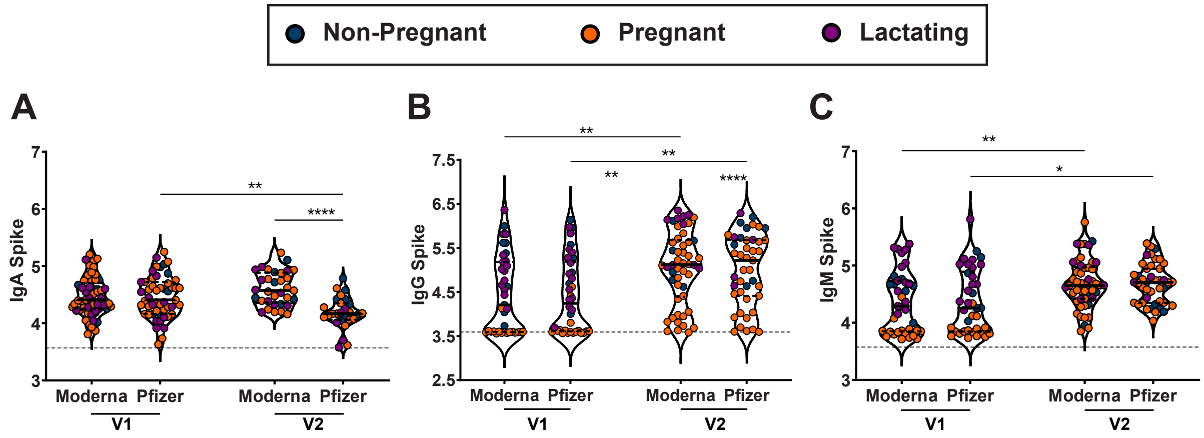


**Supplemental Figure 2. mRNA-1273 induces a greater IgA response than does BNT 162b2.** A-C. Violin plots show the log10 transformed (A) IgA Spike-, (B) IgG Spike-, and (C) IgM Spike-specific titers across V1 and V2 time points collected from non-pregnant (blue), pregnant (orange), or lactating (purple) participants receiving either mRNA-1273 (Moderna) or BNT 162b2 (Pfizer). Differences across timepoints and groups were assessed by repeated measures mixed-effects model followed by posthoc Tukey's multiple comparisons test. * p <0.05, ** p <0.01, *** p <0.001, **** p <0.0001.

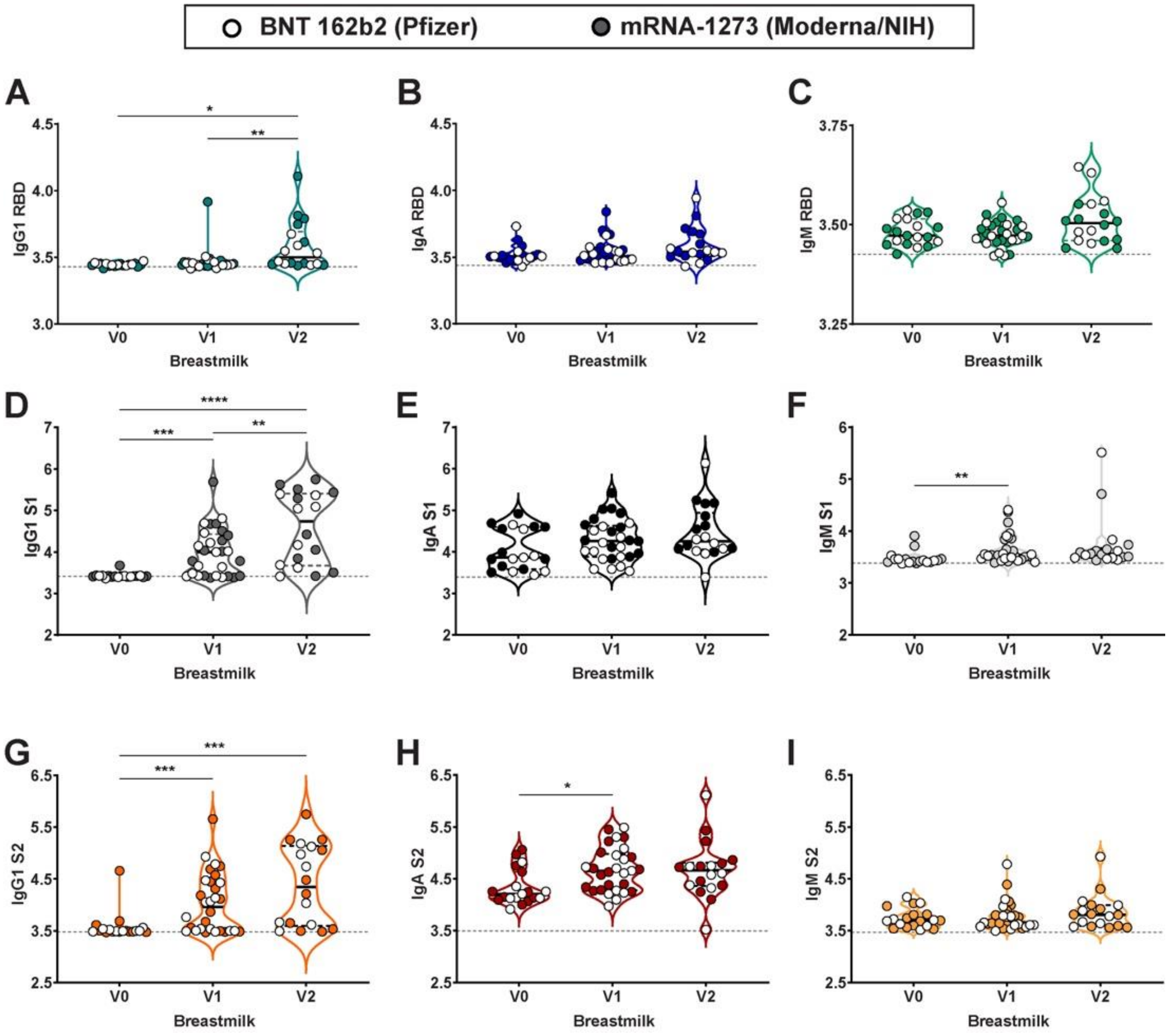


**Supplemental Figure 3. Maternal vaccination induces SARS-CoV-2-specific antibodies in breastmilk. A-I.** Violin plots show the log10 transformed **(A)** IgG1 RBD-, **(B)** IgA RBD-, and **(C)** IgM RBD-specific, **(D)** IgG1 S1-, **(E)** IgA S1-, and **(F)** IgM S1-specific, **(G)** IgG1 S2-, **(H)** IgA S2-, and **(I)** IgM S2-specific breastmilk titers across V0, V1, and V2 time points. Differences across timepoints were assessed with repeated measures mixed effects model followed by posthoc Tukey's multiple comparisons test. Participants injected with BNT 162b2 from Pfizer/BioNTech are depicted as open circles, and participants injected with mRNA-1273 from Moderna/NIH are depicted as closed circles. * p <0.05, ** p <0.01, *** p <0.001, **** p <0.0001.

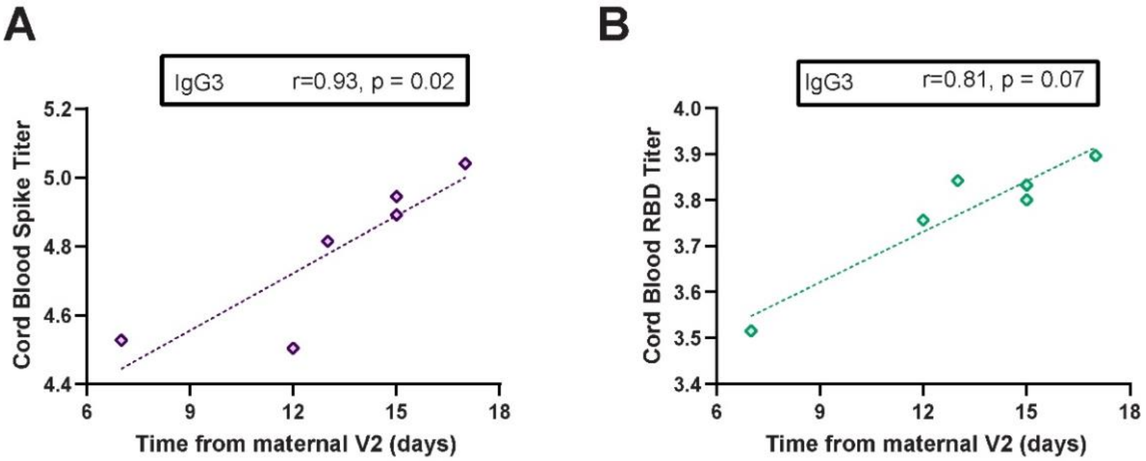


**Supplemental Figure 4. Transfer of SARS-CoV-2-specific antibodies from maternal to umbilical cord blood following maternal vaccination.**

**A-B.** The x-axis shows the time from V2 until delivery and the y-axis shows cord blood log10 transformed titer for **(A)** IgG3 Spike (purple) and **(B)** IgG3 RBD (turquoise). Significance and rho determined by Spearman's correlation test.

**Table S1. Characteristics of pregnant women with natural COVID-19 infection.**

| Characteristic | Natural COVID-19 infection in pregnancy[a] (N=37) |
|---|---|
| **Participant age**, mean (SD) | 32.5 (5.3) |
| **Race** | |
| White | 14 (39%) |
| Black | 5 (14%) |
| Asian | 1 (3%) |
| Multi-racial | 1 (3%) |
| Other | 14 (39%) |
| Unknown | 1 (3%) |
| **Ethnicity** | |
| Hispanic or Latino | 16 (44%) |
| Not Hispanic or Latino | 19 (53%) |
| Unknown/ not reported | 1 (3%) |
| **Gravidity** (including current pregnancy), median (IQR) | 2 (2, 3) |
| **Parity** (excluding current delivery), median (IQR) | 1 (0, 1) |
| **COVID Severity** | |
| Mild | 18 (50%) |
| Moderate | 11 (31%) |
| Severe | 7 (19%) |
| **Gestational age at COVID diagnosis** in weeks, median (IQR) | 30.1 (26.9, 33.8) |
| **Days from symptom onset to blood draw** in days, median (IQR) | 62.5 (39.5, 84) |

[a] Only symptomatic women/those with clinical COVID-19 included for timing of infection from symptom onset. All women had positive RT-PCR for SARS-CoV-2 by nasopharyngeal swab

**Table S2. SARS-CoV-2-specific antibody titer correlation with composite participant symptom score after vaccine dose 2.**

| **V2 timepoint antibodies** (2-6 weeks post-vaccination) | | |
|---|---|---|
| | **Spearman Rho** | **P-value** |
| ***Maternal serum*** | | |
| Spike IgG1 | 0.25 | 0.05 |
| Spike IgG3 | 0.45 | $3.0 \times 10^{-4}$ |
| RBD IgG1 | 0.29 | 0.02 |
| RBD IgG3 | 0.36 | $4.8 \times 10^{-3}$ |
| ***Breastmilk*** | | |
| Spike IgG1 | 0.49 | 0.04 |
| Spike IgG3 | 0.42 | 0.04 |
| Spike IgA | 0.42 | 0.04 |
| RBD IgG1 | 0.42 | 0.04 |
